# Breakthrough infections after post-exposure vaccination against Monkeypox

**DOI:** 10.1101/2022.08.03.22278233

**Authors:** Michael Thy, Nathan Peiffer-Smadja, Morgane Mailhe, Laura Kramer, Valentine Marie Ferré, Nadhira Houhou-Fidouh, Hassan Tarhini, Chloé Bertin, Anne-Lise Beaumont, Mathilde Garé, Diane Le Pluart, Ségolène Perrineau, Mayda Rahi, Laurène Deconinck, Bao Phung, Bastien Mollo, Marie Cortier, Mélanie Cresta, Clémentine De La Porte Des Vaux, Véronique Joly, Sylvie Lariven, Cécile Somarriba, Francois-Xavier Lescure, Charlotte Charpentier, Yazdan Yazdanpanah, Jade Ghosn

## Abstract

**Background:** A third-generation smallpox vaccine was recommended in France for individuals who had a high-risk contact with a PCR-confirmed Monkeypox patient. We aimed to describe the outcomes of high-risk contacts receiving third-generation smallpox vaccine as an early post-exposure ring vaccination (EPRV) especially tolerance and potential breakthrough infections after the first dose.

**Methods:** We performed an observational analysis of all consecutive individuals vaccinated with the IMVANEX® smallpox vaccine after a high-risk contact defined as close skin-to-skin or mucosal contact and/or indirect contact on textile or surface and/or droplets exposure defined by a contact at less than 2 meters during at least 3 hours with a PCR-confirmed Monkeypox patient.

**Results:** Between May 27^th^ and July 13^th^, 2022, 276 individuals received one dose of IMVANEX® with a median delay of 11 days [IQR 8-14] after exposure with a confirmed Monkeypox patient. Mode of exposure was droplets for 240 patients (91%), indirect contact for 189 (71%) and unprotected sexual intercourse for 146 (54%). Most of the patients were men (91%, n=250) and men who have sex with men (88%, n=233). The vaccine was well tolerated with no severe adverse event. Among the 276 vaccinated individuals, 12 (4%) had a confirmed Monkeypox breakthrough infection with no severe infection. Ten out of 12 patients developed a Monkeypox infection in the five days following vaccination and two had a breakthrough infection at 22 and 25 days.

**Conclusion:** EPRV with a third-generation smallpox vaccine was well tolerated and effective against Monkeypox but did not completely prevent breakthrough infections.

## Introduction

Monkeypox is a zoonotic disease due to an Orthopoxvirus, very similar to smallpox and first identified in 1977 in the Democratic Republic of Congo. Since the discovery of the Monkeypox virus, cases have been mainly reported during outbreaks in West and Central Africa and are thought to be related to transmission from animal to humans ^1–3^. The incubation of the disease ranges between 5 to 21 days ^3,5,9,12^. The fatality rate is poorly known, varying from 1 to 10% depending on the clade causing the disease ^4–6^. Human-to-human transmission can occur through exposure to large respiratory droplets during direct contact, through direct skin/mucosa contact with skin/mucosa lesions of an infected person, or indirectly by contact with fomites (surfaces, materials or contaminated objects) ^1,5,7^. Since May 6^th^, 2022, an outbreak has been described in western countries with non-imported cases reported by the Portuguese and British authorities and subsequently in several European countries, the United States and Canada ^8–11^. On May 19^th^, 2022, a first case of Monkeypox was confirmed in France. On July 12^th^, 2022, the French national agency of public health (Santé publique France) declared 912 confirmed cases of Monkeypox with no direct link to people returning from endemic areas ^11^. This outbreak has distinct characteristics than previously reported outbreaks. Indeed, the vast majority of infections occurs in men having sexual relationships with men (MSM) and transsexuals, with a high prevalence of pharyngeal, anal and genital lesions ^9^.

There is no specific vaccine to prevent Monkeypox but a third-generation vaccine against smallpox has been validated and is available for the indication of active immunization against smallpox ^13–16^. Stocks exist in numerous countries for the potential bioterrorism threat of smallpox. This vaccine is a non-replicating live vaccine containing a live modified form of the vaccinia Ankara virus (MVA), produced by Bavarian Nordik and authorized since 2013 in Europe under the trade name IMVANEX® ^8,17^. Vaccination after a Monkeypox exposure may help to prevent the onset of the disease or make it less severe ^18–20^. In the context of the current outbreak, early post-exposure ring vaccination (EPRV) with smallpox vaccine (off-label use) has been recommended by WHO, USA Centers for Disease Control & Prevention (CDC) and France to reduce symptoms of Monkeypox disease and to limit transmission ^8,21–24^. In France, the French high authority of health (HAS) has recommended the implementation of a reactive vaccination strategy in early post-exposure with the third-generation vaccine administered in 2 doses spaced 28 days apart, the first dose being ideally administered within 4 days after the high-risk exposure with a PCR-confirmed Monkeypox patient and no longer than 14 days after the high-risk exposure ^23^. The aim of this study was to describe the outcomes of high-risk contacts receiving third-generation smallpox vaccine and in particular tolerance of the vaccine and potential breakthrough infections within the 28 days after the first dose of vaccine.

## Methods

### Study population

In this study, we followed all consecutive patients who were vaccinated by the third-generation smallpox vaccine (IMVANEX®) as part of the EPRV strategy in Bichat Claude Bernard University Hospital from May 27^th^ to July 13^th^, 2022. The vaccine was administered according to French guidelines as part of care ^23^. Vaccinated persons were those exposed to a PCR-confirmed case of Monkeypox; exposure was defined according to French high public health council (HCSP) guidelines ^25^. It included direct skin-to-skin or mucosal contact including sexual intercourse with a confirmed Monkeypox patient, indirect contact with a confirmed Monkeypox patient through fomites (textiles or surfaces) and/or droplets exposure defined by a contact at less than 2 meters during at least 3 hours with a confirmed Monkeypox patient. Contra-indications to vaccination included hypersensibility and allergy to vaccine components (chicken proteins, benzonase, gentamicin, ciprofloxacin) ^17,26^. All participants received at least one dose of the IMVANEX® vaccine and we assessed the proportion of participants breakthrough infection within the 28 days after the first vaccine dose. All vaccinated individuals provided written informed consent for the collection and analysis of data. The local review board approved the study.

### Data collection

Demographic data, underlying medical conditions including immunosuppression (immune deficiency, HIV, immunosuppressive drugs including corticosteroids), history of smallpox vaccination and mode of exposure with a confirmed Monkeypox case were collected. Patients had a clinical examination to look for signs of Monkeypox infection including systemic symptoms and cutaneous and/or mucous lesions. If signs compatible with Monkeypox infection were reported, the data were collected and PCR for Monkeypox virus was performed on the lesions with at least one and up to three samples collected: throat swab, skin swab, EDTA blood sample. The skin swab was rubbed against all detected skin lesions and a pustule was popped if possible, in order to increase sample sensitivity. After a heat inactivation step (12 minutes at 70°C), nucleic acids were extracted using MagNA Pure LC 2.0 Instrument (Roche, Meylan, France). Monkeypox virus specific real-time PCR assay validated by both CDC and French *Orthopoxvirus* national reference center was performed ^27^. An exogenous internal extraction and amplification control (IEAC) was added to each sample before extraction (Simplexa™ Extraction and Amplification Control Set, DiaSorin, Saluggia, Italy) ^28^. Negative controls were added to each extraction batch and a positive control was tested in each PCR run. Confirmed Monkeypox cases were defined as individuals with positive PCR assay on any kind of sample.

If the PCR for Monkeypox were negative, vaccination was carried out after 48 hours without symptoms.Vaccinated contacts were questioned by e-mail about side effects or symptoms since vaccination on July 13^th^, 2022. Suspected Monkeypox disease cases after vaccination were called to the hospital and at least two samples were collected for PCR: one oropharyngeal and one on all the cutaneous and mucous lesions.

Results are expressed in medians and interquartile [IQR] as well as absolute numbers and proportions for categorical data.

## Results

Between May 27^th^ to July 13^th^, 2022, 284 patients presented for EPRV. Among them, 31 (11%) patients were symptomatic on the day of the appointment with a median delay between the Monkeypox exposure and symptoms of 7 [IQR 4-8] days. Among the 31 patients, 23 had a negative PCR and were vaccinated. The eight other patients either had a positive PCR on skin lesions or were considered clinically highly suspicious of Monkeypox infection.

In total, 276 individuals had one dose of IMVANEX® with a median [IQR] delay of 11 [8-14] days after exposure with a confirmed Monkeypox patient. A flow chart is displayed on Fig. 1. Mode of exposure was droplets for 240 patients (91%), indirect contact for 189 (71%) patients and unprotected sexual intercourse for 146 patients (53.7%). Most of the patients were men (91%, n=250) and men who have sex with men (MSM) (88%, n=233) and were born in France (87%, n=177). Twenty-nine patients (11%) patients had a history of vaccination against smallpox between 1955 to 1981 (27 patients had had one injection and 2 patients had had two injections). The main characteristics of the vaccinated individuals are shown on Table 1.

**Figure 1:**
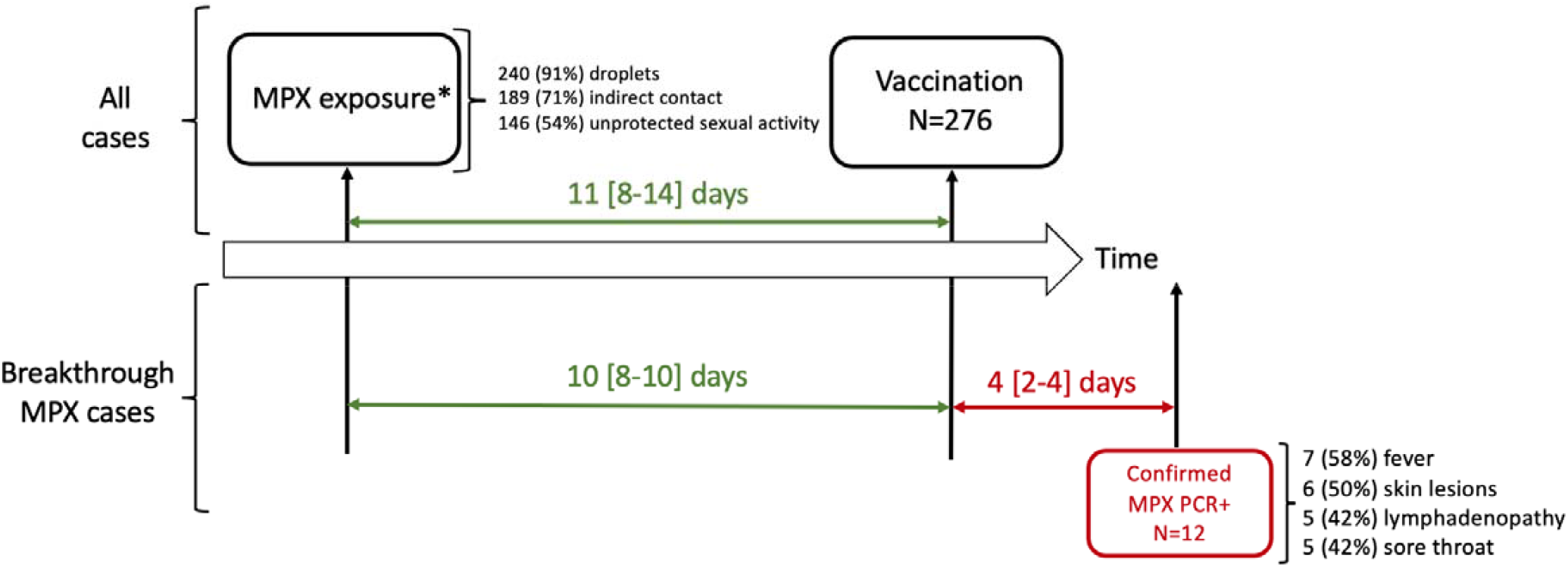
Flow chart of early post-exposure vaccination against Monkeypox. MPX: Monkeypox *MPX exposure: direct skin-to-skin or mucosal contact including sexual intercourse with a -confirmed monkeypox patient, indirect contact with a confirmed monkeypox patient through fomites (textiles or surfaces) and/or droplets exposure defined by a contact at less than 2 meters during at least 3 hours with a confirmed monkeypox patient

**Table 1:** Main characteristics of the early post-exposure ring vaccinated population and comparison between who developed symptoms or not after vaccination.

|  | Total | Breakthrough<br>Monkeypox<br>infections* |
| --- | --- | --- |
| n | 276 | 12 |
| Age (median [IQR]) | 19.0 [14.0-25.0] | 24.0 [15.8-26.8] |
| Male (%) | 250 (90.6) | 11 (91.7) |
| Born in France (%) | 177 (87.2) | 8 (88.9) |
| MSM (%) | 233 (88.3) | 11 (91.7) |
| Number of sexual partners during past month (median [IQR]) | 8.0 [3-13] | 11.0 [4-12.5] |
| Number of STIs during past year (median [IQR]) | 1.5 [1-2] | 1.5 [1-2] |
| Chemsex user (%) | 75 (28.6) | 4 (33.3) |
| Domestic animal (%) | 57 (21.0) | 2 (18.2) |
| Past medical history (%) | 73 (26.7) | 3 (25.0) |
| Cancer or blood disease (%) | 2 (0.7) | 0 (0.0) |
| HIV (%) | 38 (13.9) | 4 (33.3) |
| Immunodepressed (%) | 3 (1.1) | 0 (0.0) |
| Past history of STIs (%) | 144 (61.5) | 8 (72.7) |
| PreP user (%) | 138 (51.3) | 5 (41.7) |
| Past history of smallpox vaccination (%) | 29 (10.5) | 2 (16.7) |
| Symptomatic before vaccination (%) | 23 (8.3) | 0 (0.0) |
| Fever before vaccination (%) | 4 (1.4) | 0 (0.0) |
| Skin lesions before vaccination (%) | 16 (5.8) | 0 (0.0) |
| Healthcare worker (%) | 30 (10.9) | 2 (16.7) |
| Travel during past month (%) | 96 (35.8) | 3 (25.0) |
| Exposure with confirmed Monkeypox by PCR (%) | 259 (98.5) | 7 (100.0) |
| Relationship with confirmed Monkeypox (%) |  |  |
| Occasional partner | 132 (53.2) | 3 (42.9) |
| Friend | 46 (18.5) | 3 (42.9) |
| Permanent partner | 17 (6.9) | 0 (0.0) |
| Patient | 14 (5.6) | 1 (14.3) |
| Room mate | 10 (4.0) | 0 (0.0) |
| Family | 11 (4.4) | 0 (0.0) |
| Droplets exposure (%) | 240 (90.6) | 11 (91.7) |
| Indirect exposure (%) | 189 (71.1) | 10 (83.3) |
| Unprotected sexual intercourse (%) | 146 (53.7) | 5 (41.7) |

**Legends:** \* confirmed with PCR
MSM: men having sex with men; STIs: sexually transmitted infections; HIV: human immunodeficiency virus; PreP : HIV pre-exposure prophylaxis

**Table 2:** Details of side effects and symptoms after early post-exposure ring vaccination.

|  | Total | Breakthrough<br>Monkeypox<br>infections* |
| --- | --- | --- |
| n | 276 | 12 |
| **Side effects of vaccination (%)** | **49 (50.0)** | **0 (0.0)** |
| Local pain (%) | 43 (46.2) | 0 (0.0) |
| Fatigue (%) | 13 (15.5) | 0 (0.0) |
| Delay between exposure and symptoms (median [IQR]) |  | 12.0 [9.3-15.3] |
| Delay between vaccination and symptoms (median [IQR]) |  | 4.0 [1.8-4.3] |
| **Positive Monkeypox PCR (%)** | **12 (54.5)** | **12 (100.0)** |
| **Symptoms** |  |  |
| Fever after vaccination (%) | 11 (4.7) | 7 (58.3) |
| Body aches after vaccination (%) | 4 (1.7) | 2 (16.7) |
| Lymphadenopathy after vaccination (%) | 5 (2.1) | 5 (41.7) |
| Sore throat after vaccination (%) | 6 (2.6) | 5 (41.7) |
| Cough after vaccination (%) | 1 (0.4) | 1 (8.3) |
| Headaches after vaccination (%) | 1 (0.4) | 0 (0.0) |
| **Skin lesions** |  |  |
| Anal lesions (%) | 6 (2.6) | 5 (41.7) |
| Rectitis (%) | 3 (1.3) | 2 (16.7) |
| Face lesions (%) | 3 (1.3) | 2 (16.7) |
| Trunk lesions (%) | 6 (2.6) | 4 (33.3) |
| Limbs lesions (%) | 2 (0.9) | 2 (16.7) |
| Genital lesions (%) | 2 (0.9) | 1 (8.3) |
| Associated STI (%) | 4 (1.6) | 1 (8.3) |
| **Length of follow-up after vaccination (median [IQR])** | **5.5 [1.0-20.0]** | **13.5 [5.8-19.3]** |

Legends: \* confirmed with PCR STI: sexually transmitted infection

Among 101 (37%) individuals who answered to the e-questionnaire regarding vaccine tolerance, 45 (45%) reported local pain and 13 (13%) fatigue during a median of 4 [3-8] days after vaccination. No fever or other systemic symptoms were described. One vaccinated patient was allergic to ciprofloxacin but no side effects were described after vaccination.

Among the 276 vaccinated individuals, 12 (4%) had a confirmed Monkeypox breakthrough infection confirmed with ten positive PCR on skin lesions and/or seven positive oropharyngeal PCR. They had a median age of 24 [16-27] years old, eleven out of the 12 cases were MSM (92%) with a median number of 11 [4-12] sexual partners during the last month, a median of 2 [1-2] STIs during the last year and 4 (33%) were Chemsex users. One breakthrough infection occurred in a healthcare worker by accidental occupational direct inoculation with a needle. The other modes of contamination were exposure through droplets for 11 patients, indirect exposure for 10 patients and unprotected sexual intercourse for five patients.

Among these 12 Monkeypox breakthrough infections, the median delay between the exposure to a confirmed case of Monkeypox and the vaccine was 10 [8-10] days with a range from 0 to 12 days. The median delay between the vaccine and the first symptoms of Monkeypox infection was 4 [2-4], ranging from 1 to 25 days. Ten out of 12 patients developed a Monkeypox infection in the 5 days following vaccination. Among the 12 breakthrough infections, the two patients who were symptomatic later than 5 days after vaccination (22 and 25 days) were the only ones with a pet animal. The delays between exposure, vaccination and confirmed Monkeypox infection are displayed on Fig. 2.

**Figure 2:**
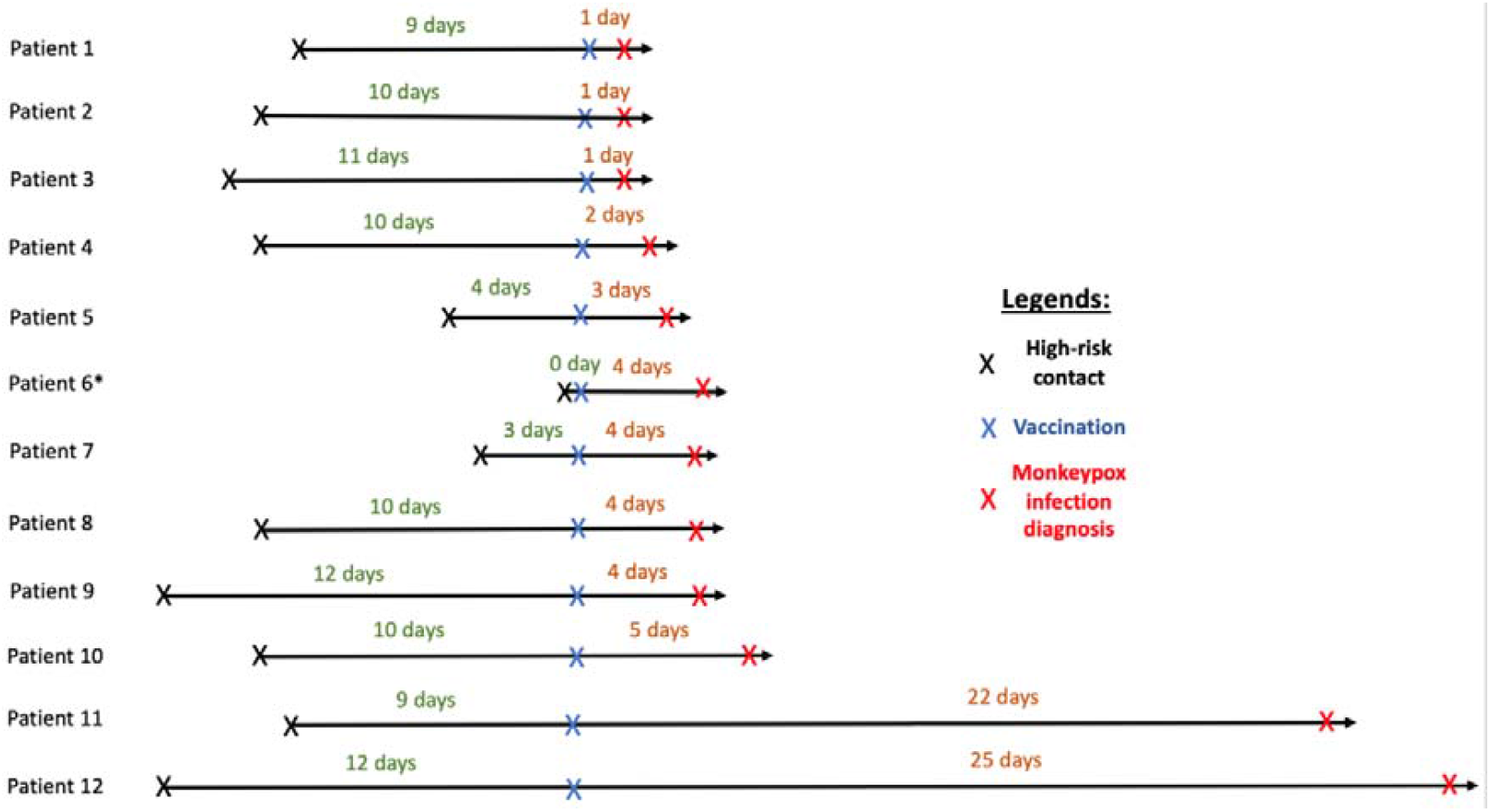
Delay between exposure, vaccination and confirmed Monkeypox infection in the 12 breakthrough infections. *Patient 6: Direct inoculation with a percutaneous needlestick

The systemic symptoms of Monkeypox infection included fever for seven (58%) patients, lymphadenopathy for five (42%), sore throat for five (42%), body aches for two (17%) and cough for one (8%). Six (50%) patients developed a median of 2 [1-3] skin lesions. Affected areas included the anal region (n=5 patients) including two with rectitis, the trunk (n=4), the face (n=2), the limbs (n=2), the genital area (n=1). There were no severe infections requiring hospitalization. The median [IQR] Ct of the positive PCR was 24 [22-28] on skin lesions and 30 [28-34] on oropharyngeal samples.

Ten other patients declared skin lesions compatible with a Monkeypox infection on the questionnaire. Six had a negative PCR and four declined to come back to the center for PCR testing.

Among the patients who did not develop Monkeypox and had no history of smallpox vaccination, the 2^nd^ dose of the vaccine was performed after a median of 29 [28-33] days.

## Discussion

This study assessed the outcomes of high-risk contacts receiving one dose of the third-generation vaccine IMVANEX® used in a strategy of early post-exposure ring vaccination (EPRV). The median delay between exposure and vaccination was 11 [8-14] days post exposure to Monkeypox. In this study the vaccine was well tolerated with no serious adverse events. Regarding efficacy, 4% (12/276) of the individuals had a PCR-confirmed breakthrough Monkeypox infection after a confirmed high-risk exposure to Monkeypox.

The 12 patients who developed a confirmed Monkeypox disease were vaccinated after a median delay of 10 [8-10] post exposure; 50% of cases with breakthrough infections having received the vaccination after 13 days and 75% after eight days. This relatively long delay may explain early breakthrough infections as the ideal timing for vaccination is thought to be in the four days following exposure. Indeed, the incubation of the Monkeypox virus has been described to range from 5 to 21 days and delayed vaccination may be too late to prevent the disease in some patients. This delay underlines the constraints that exist on the field to implement an EPRV strategy during an outbreak. Especially during an outbreak of an emerging infectious disease, procedures of contact tracing take time and may delay the access to vaccination after confirmed exposure. Early diagnosis of cases and efficient contact tracing are key public health interventions to control an epidemic in general and they may also allow a more successful and efficacious EPRV strategy.

Ten out of the 12 breakthrough infections occurred in the five days following vaccination. The two other patients had a Monkeypox infection 22 and 25 days after vaccination. Post-exposure prophylaxis (PEP) was used in 2018 and 2019 in the UK ^29^. In 2018, 3 Monkeypox cases were declared among 154 contacts (including 147 healthcare workers). Among them, 131 received PEP (126 healthcare workers), and one secondary case occurred on a healthcare worker who received one dose of IMVANEX® vaccine after exposure on day 6 post-exposure or 12 days pre-illness. In 2019, 17/18 contacts (including children) were vaccinated after contact with one imported case but no Monkeypox case was declared after vaccination^29^. A phase 1 study found that with a single-dose of third-generation MVA smallpox vaccine in humans, the peak of antibody titer was reached at day 14 with a decrease of antibodies from day 14 to the second dose of vaccine ^30^. Therefore, the fact that 10 out of 12 cases occurred before 5 days after vaccination is not surprising. We were more surprised by the 2 cases in whom the infection occurred after 20 days. Whether the decrease of antibodies from day 14 may explain the late breakthrough infections, especially if the patient had a new exposure to the Monkeypox virus, it cannot be confirmed based on our data but could be a hypothesis. We did not find any new exposure for these two patients but they were the only patients out of the 12 who possessed a pet animal. The pet could theoretically be a reason for persistent exposure in these patients as domestic animals such as cats and dogs can be infected with Monkeypox virus ^31,32^. If confirmed, the hypothesis of a reinfection may strengthen the importance of the second dose of vaccine on Day 28 and research around this question should be reinforced.

Although we should be cautious because of the limited number of cases enrolled in this study, it is important to note that none of the 12 breakthrough infections after vaccination was severe or complicated. The few data available on the severity of the Monkeypox infection after a third-generation smallpox vaccine show that after challenge with Monkeypox virus, the vaccinated animals were healthy and asymptomatic compared to unimmunized animals which developed more than 500 pustular skin lesions and became gravely ill or died ^16,33–36^.

In recent guidelines, the EPRV strategy has been associated with a strategy of pre-exposure vaccination of the patients with multiple sexual casual partners (including MSM, transgenders, sex workers), regardless of a documented exposure. Past data from Africa suggest that the smallpox vaccine is at least 85% effective in preventing Monkeypox when given before exposure to Monkeypox ^1,14^. The two strategies, EPRV and pre-exposure vaccination, should be complementary to fight against the spread of Monkeypox in the general population.

This study had limitations. First, the design of the study was not adapted for a precise evaluation of the tolerance and efficacy of a vaccine which requires clinical trials or comparisons between exposed individuals who are or are not vaccinated. However, it seemed important to describe breakthrough infections as part of the EPRV strategy. Second, no systematic follow-up clinical visit was implemented after vaccination and four individuals reported symptoms compatible with a breakthrough infection but declined to come back to be tested. Third, the monocentric design could limit external validity but this center was the only vaccination center in France until June 14^th^, 2022 (for almost a month). Fourth, the measure of the immunity and in particular antibodies level induced by vaccination was not performed in this study. Evidence on the effectiveness of an EPRV strategy after Monkeypox exposure is limited but this study confirms its potential safety and efficacy.

## Conclusion

In this cohort of 276 individuals vaccinated with a third-generation smallpox vaccine after a high-risk contact with a PCR-confirmed Monkeypox patient, only 12 (4%) breakthrough infections were observed. None of the infections were severe or complicated. These data need to be confirmed in further clinical and immunological studies.

## Data Availability

All data produced in the present study are available upon reasonable request to the authors

## Acknowledgments

The authors thank Santé publique France (SPF) and Agence Régionale de Santé (ARS).

